# Studying social anxiety without triggering it: Establishing an age-controlled cohort of social media users for observational studies

**DOI:** 10.1101/2023.12.11.23299645

**Authors:** Ana Lucia Schmidt, Karen O’Connor, Graciela Gonzalez Hernandez, Raul Rodriguez-Esteban

## Abstract

**Background:** Patients of certain diseases are less likely to approach the healthcare system but remain active in social media. Young Social Anxiety Disorder (SAD) patients, in particular, are a hard-to-reach population due to disease symptomatology, unmet need and age-related barriers, which makes obtaining first-hand access to patient perspectives challenging.

**Objective:** To create a curated cohort of patients from social media that report their age in the range of 13 to 25 years old and confirm having a SAD diagnosis or having received therapy for SAD, and to assess the value of the content posted by these users for observational studies of SAD.

**Methods:** We collected 535k posts by 118k Reddit users from the r/SocialAnxiety subreddit. We then developed precise regular expressions to extract age, diagnosis and therapy mentions. We manually annotated the full set of expressions extracted and double-annotated 5% of the age mentions and 10% of the diagnosis and therapy mentions. Using similar methodology, we identified mentions of comorbidities and substance use.

**Results:** Our validated cohort includes 37,073 posts by 1,102 users that meet the inclusion criteria. The age, diagnosis, and therapy mention detection had a precision of 68%, 31%, and 44%, respectively, with an inter-annotator agreement of 0.96, 0.96, and 0.78. Sixty-one percent of the users in the cohort report having one or more comorbidities on top of their SAD diagnosis (Fleiss’s Kappa=0.79) and 13% report a concerning use of drugs or alcohol (Fleiss’s Kappa=0.87). We compared the characteristics of our social media cohort to the published literature on SAD.

**Conclusions:** Patients with SAD post actively on Reddit and their perspectives can be captured and studied directly from these data. Extracting age, therapy, substance abuse and comorbidities (and potentially other patient data) can address realworld data source biases. Thus, social media is a valuable source to create cohorts of hard-to-reach patient populations that may not enter the healthcare system.

## 1 Introduction

Social Anxiety Disorder (SAD) is characterized by marked and persistent fear of one or more social situations in which the person is exposed to unfamiliar people or to possible scrutiny by others^1,2^ and has an estimated prevalence ranging from 4%^3^ to 36%^4^. It has an early onset, with 50% of individuals experiencing symptoms by age 13 and 90% by age 23, and frequently persists into adulthood and even old age^5^. For patients with SAD, exposure to social or performance situations invariably triggers anxiety, which can lead to avoidance of the specific trigger and significant impact in their everyday functioning and their quality of life. SAD is associated with increased presence of comorbid mental disorders^6^, substantial impairment across multiple domains^7^, lower health-related quality of life^8^ and issues with substance abuse^9^. The majority of individuals with SAD have additional psychiatric disorders^6,10^, primarily other anxiety disorders^6^, Major Depressive Disorder (MDD)^11^, mood disorders^12^, eating disorders^13,14^ and substance abuse problems^9,15^.

Despite SAD being prevalent among adolescents, it paradoxically stands as one of the least recognized and treated pediatric disorders. The vast majority of research on the condition has focused on adults^16^. Most approaches to studying adolescents with SAD usually rely on retrospective studies in which adults recall their experiences^7,11,17,18^. These studies suffer from known limitations such as reporting bias^19^, recall bias^20^, negativity bias^21^ and post-event processing issues^22^. While there have been studies focused on getting information directly from adolescent patients^23,9^, obtaining clinically relevant insights from adolescents, such as patient-reported outcomes, is complicated by challenges with recruitment and patient retention^24^. Best practices for adolescent recruitment emphasize the need to employ a wide variety of recruitment methods, such as text messages, social media, and word of mouth^25^, and a focused and judicious strategy to reduce the participant’s burdens^26^ to maximize cohort retention. This is particularly aggravated in the case of SAD, as the disease is the main obstacle to participation with patients reluctant to interact with researchers, both in-person and virtually. On the other hand, socially anxious individuals engage more online, even to a problematic degree, as a diversion and to seek out social support^27,28,17^. Thus, Social Media Listening (SML), whereby publicly available data posted by patients is collected to extract insights, has the potential to be a complementary study approach that allows systematic access to this hard-to-reach populations while eliminating the burden of recruitment and personal interaction.

SML is the systematic use of automatic and manual methods that derive relevant insights from social media data for observational studies^29^. SML has been already applied for research into mental disorders^30,31,32,33,34^ such as depression^35,36,33^, suicide^37,38^, ADHD^39,33^ and anxiety^40,33^. However, most SML studies related to mental health use an aggregate approach, studying the language and characteristics of large quantities of social media data without delving into specific patient cohorts nor validating the presence of a diagnosis. Cohort studies are a type of longitudinal study in which research is done on a group of participants who share a common characteristic over some time. Retrospective cohort studies with SML data have been done in the past for cases such as Inflammatory Bowel Disease^41^ and pregnancy^42^, but we could not identify any SML study of mental disorders that use this approach.

In this study, we chose a retrospective cohort study approach with diagnosis validation because it enables rigorous analysis from social media data by systematically gathering publicly-shared patient profile-relevant insights. These data, collected from patients 13 to 25 years old, should be a valuable tool for future studies of SAD impairment in daily functioning, its impacts in quality of life, and barriers to diagnosis and treatment.

The rest of this paper describes how we automatically extracted and validated a cohort of Reddit users active in the r/SocialAnxiety subreddit who report being between 13 to 25 years old and either have a SAD diagnosis or declare having received therapy. We then outline the extraction pipeline for identifying reports of psychiatric comorbidities, particularly substance use disorders, and demonstrate the potential value of the extracted data by comparing reporting rates to those published in the relevant literature.

## 2 Materials and Methods

### 2.1 Data Collection

The data was collected from Reddit, a social news website and forum where content, grouped in topic-specific forums known as subreddits, is socially curated and promoted by site members through voting. Reddit member registration is free and anonymous and it is required to use the site’s basic features, although no account is required to read the content as it is made publicly available. r/SocialAnxiety is a support subreddit about SAD. Like all subreddits, r/SocialAnxiety is moderated and has a set of rules users must follow to keep discussions cordial and on topic.

All posts from this subreddit were retrieved using the Pushshift API on March 23, 2022. Pushshift was a platform, no longer active, for the mining, analysis, and archiving of social media data that collected Reddit data and made it available to researchers from 2015 until 2023^43^. The data is still available and can be found at Academic Torrents^44^. The dataset consists of all the posts on the r/SocialAnxiety subreddit available at the time of download. Posts can be of two types:

- A thread head, which is the first post of a new thread within the subreddit. These posts have a title that labels the new thread.
- A comment, which is any post made in an existing thread.

The dataset elements consist of post ID, author, title of the thread (if the post is a thread head), post text and timestamp. Thread heads have a mean length of 898 (sd=964) characters and comments have a mean length of 254 (sd=364) characters. Table 1 contains an overview of the downloaded data and Table 2 contains example posts.

**Table 1:** Breakdown of the collected data, all from the r/SocialAnxiety subreddit, with posts spanning from January 10, 2015 through March 23, 2022.

|  | # |
| --- | --- |
| **Threads** | 91,881 |
| **Users** | 118,040 |
| **Posts** | 535,544 |
| Thread Heads | 91,881 |
| Comments | 443,663 |

**Table 2:** Examples of posts.

| Post | Post Type |
| --- | --- |
| **Alcohol to feel better?** I'm a 25 year old guy that suffers from social/general anxiety. I find myself drinking alcohol before going to work to numb down my anxiety. I know it's not right or a safe thing to do, but it helps me tremendously. Anyone else here drink a little before work or before going anywhere to lower your anxiety? | Thread Head |
| **How can I get over my social anxiety?** its ruining my life. I have tried CBT therapy, I don't want to take medication as I have mild dystonia already and don't fancy full blown dystonia so I am kind of at a loss, I get constant dizziness, swaying, sickness, crushing pains in my head and face, hyperawareness of facial movements and body posturing when speaking to people its a nightmare now. | Thread Head |
| Hello! I am a 21 F with depression and social anxiety! I'm still trying to beat both, so good for you for getting rid of social anxiety!!!! If I fit your qualifications, I'd love to listen whenever we are both free to chat. :) | Comment |
| I'm 23 and have been diagnosed with social anxiety, depression and OCD. I've recently started medication which has been helping a lot. I found speaking foreign languages has been a huge help. If I make a faux pas or make a fool of myself, the other person can chalk it up to me just not being fully fluent in the language yet. Or at least that's how I imagine it in my head. It's a bit nerve wracking at first, but after you have one or two conversations you can get in the swing of it. In addition to my two native languages, I speak 6 others at various levels. I also love solo travel, writing, reading, hiking, stationery (idk why, but it calms me) and music. | Comment |
| I'm 23 and have been diagnosed with social anxiety, depression and OCD. I've recently started medication which has been helping I have, I'm going to therapy and will continue to. Thank you, I appreciate it! | Comment |

### 2.2 Cohort Creation

From all the users in the r/SocialAnxiety subreddit, we focused on the extraction of a cohort of users that:

1. report being 13 to 25 years old at the time the posts were made, and
2. report having social anxiety by expressing that they have been diagnosed or that have received therapy for SAD.

All the posts were processed in a series of steps, as seen in Figure 1, whereby relevant data was extracted and then manually annotated by 2 different annotators. The guidelines relevant to the different annotation steps can be found in the Supporting Information.

**Figure 1:**
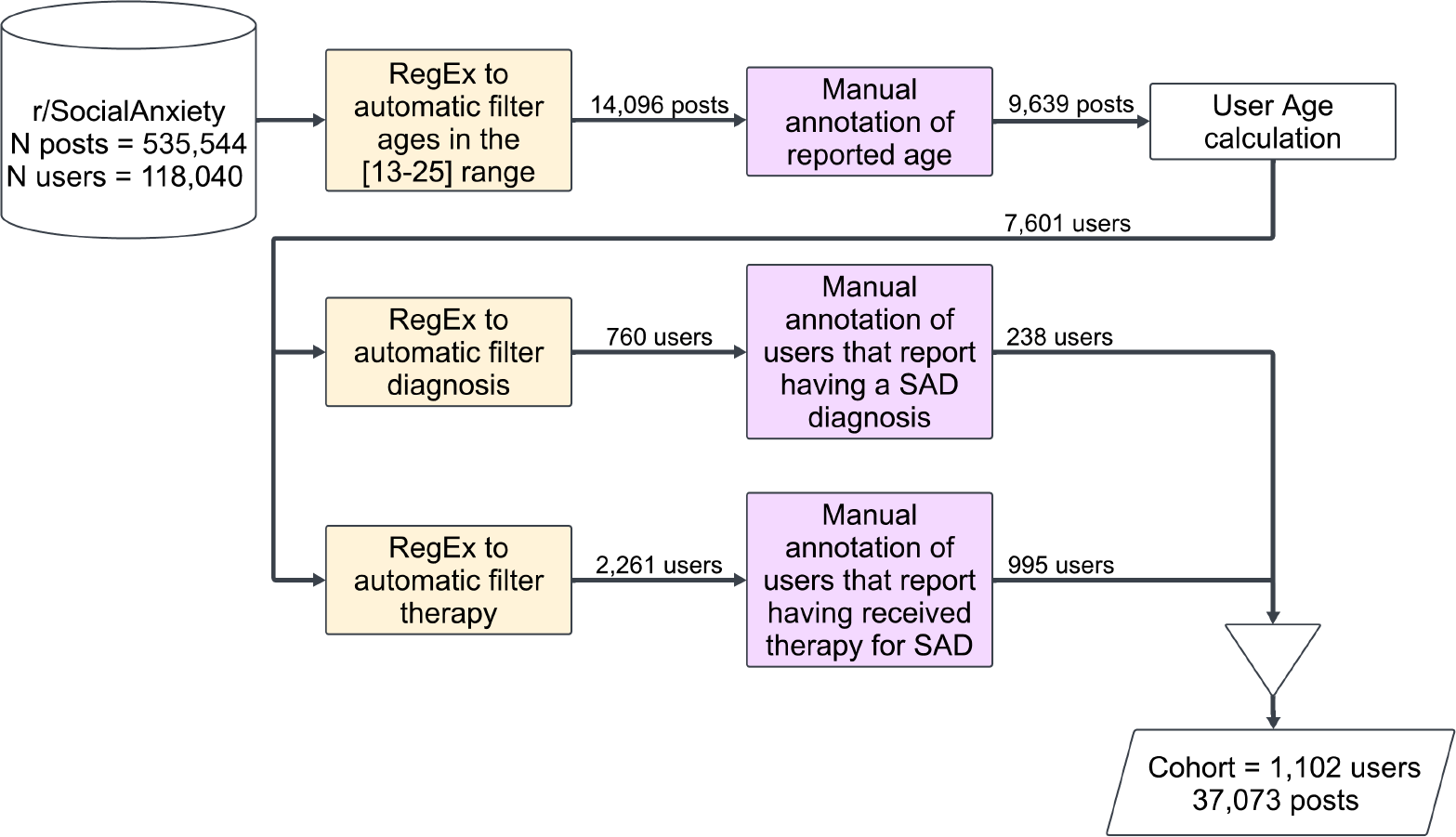
Diagram of the cohort creation pipeline.

This allowed us to generate a cohort that meets the inclusion criteria.

#### 2.2.1 Age Detection

As seen in Figure 1, the first step of the cohort creation pipeline consists of selecting all users who report being 13 to 25 years old. To do this, we first cleaned the posts by removing all URLs and replaced written numbers in the 1 to 99 range for their equivalent 2-digit representation. We then removed all posts that do not have numbers in the 13 to 25 range. Finally, we used regular expressions to automatically filter sentences that do not contain a self-reported age of the user. Some of these regular expressions identify numbers that refer to time (hours, minutes, seconds, days, etc.), dosage (pills, capsule, vial, drops, etc.), and other common expressions that use 2-digit numbers that are not related to age (e.g. money, percentages, COVID-19, etc.). The regular expressions used to filter non-relevant posts can be found in the Supporting Information.

We proceeded to manually annotate the 14, 096 posts (from 10, 670 users) that passed the automatic filters. The annotators labeled the posts with the reported age or NA if no age could be determined. A random sample of 5% was double annotated to calculate the inter-annotator agreement.

#### 2.2.2 Age Calculation

Because age annotation was done on individual posts, a user can be associated to multiple ages over time on r/SocialAnxiety. Thus, we estimated a reference age for each user. For each user, all age annotations were used to calculate their birth year. When a user had multiple age annotations, the median birth-year value of the set was used.

For the analysis, the reference age of the user was the year the post was written minus the estimated birth year. Given that user activity duration has a long-tail distribution in which the median is 16 days, the mean is 181 days and the standard deviation is 323 days, we believe this reference age is a fair approximation of the users’ ages for our analyses.

#### 2.2.3 Detection of SAD diagnosis with different levels of certainty

After extracting a group of users that reported being in the correct age range of [13-25], we then proceeded to ensure the users reported having Social Anxiety Disorder. This was done in two ways:

- by searching for a reported SAD diagnosis, and
- by searching reports of therapy for SAD.

To confirm the presence of a diagnosis we collected, for each user, a series of chronologically sorted sentences that included the substring *“diagno*.*”* Of the 7, 601 users analyzed only 760 users had matches, which were then manually labeled: ‘1’ in the case the of a SAD diagnosis report, ‘0’ otherwise. A 10% sample was double annotated to calculate the inter-annotator agreement.

Similarly, we confirmed the users’ exposure to therapy for SAD by extracting, for each user, a series of chronologically sorted sentences that matched a set *t* of regular expressions related to therapy, those being, *t = {“therapy”, “counseling”, “session”, “\\bappointment“, “treat(ed*|*ment)”, “psychologist”, “psychotherapist”, “shrink”, “therapist”, “doctor”, “psychoanalyst”, “clinician”}*, and then annotating those sentences. This approach assumes that a user who is receiving, or has received in the past, professional help for SAD most likely has been diagnosed. While this assumption does not provide the same level of certainty as the detection of a reported diagnosis, we believe it is a fair approximation to the communication style of patients online.

Of the 7, 601 users analyzed, only 2, 261 matched therapy expressions and were manually labeled: ‘1’ if the user reported attending or having attended therapy for SAD, or ‘0’ otherwise. Like the diagnosis annotation, a 10% sample was double annotated to calculate inter-annotator agreement.

Once the two sets of users were annotated, we merged them while keeping track of the type of diagnosis evidence (direct or indirect) found in the text.

### 2.3 Detection of Comorbidities and Substance Abuse Disorders

We annotated comorbidities and substance abuse (alcohol and/or drugs) as reported in Reddit by the subjects in our cohort. This was done similarly to the previous steps, that is, utilizing regular expressions followed by manual annotation. We created 2 filters based on regular expressions: one for comorbidities and one for substance abuse. The regular expressions were generated and refined after multiple iterations over the data and comprised mental disorders, their linguistic variations, abbreviations, and substance abuse-related keywords. They included conditions such as general anxiety disorder, panic disorder, bipolar disorder, schizoaffective disorder, and obsessive-compulsive disorder, among others, as well as substance abuse-related keywords such as “alcoholic,” “addiction,” “drunk,” “drugs” and “weed.” The complete list of regular expressions and annotation guidelines is provided as Supplementary Material.

After passing the automatic filters, we created 2 sets of chronologically sorted sentences grouped by user, one for comorbidities and one for substance abuse, that were fully labeled by one annotator, and 10% of each set was double annotated to measure agreement. Multi-label annotation was permitted.

In the case of comorbidities, the annotators had to label all the reported comorbidities or NA if none was present. While a comprehensive list was given to the annotators, they were also provided the option to annotate mental disorders outside the given set. It is worth noting that, unlike the labeling of SAD diagnosis, the labeling of reported comorbidities was less strict in terms of what was considered a positive report. Thus, cases of users who report having a given comorbidity have the same validity as users who report having a diagnosis for said comorbidity.

In the substance abuse annotation, the labels to annotate were:

- ‘Not concerning’: the user reports having tried alcohol and/or drugs in the past, or describes a normal drinking behavior or normal recreational use of legal drugs.
- ‘Concerning use of alcohol’: the user reports drinking to cope with SAD, but does not describe a behavior that seems to indicate addiction.
- ‘Concerning use of drugs’: the user reports consuming recreational drugs to cope with SAD, but does not describe a behavior that seems to indicate addiction.
- ‘Addiction Alcohol’: the user reports having an addiction problem with alcohol or having had an addiction problem in the past, or describes a behavior that indicates addiction.
- ‘Addiction Drugs’: the user reports having an addiction problem to drugs or having had an addiction problem in the past, or describes a behavior that indicates addiction.
- ‘NA’: the user talks about alcohol and/or drugs but not about their own relationship to drugs and alcohol (e.g. they talk about other people, hypothetical situations, etc).
- ‘Other’: the user reports having other kinds of addictions the annotator considers worth labeling beyond the set of labels given.

More details about the annotation, the complete annotation guidelines, as well as the regular expressions used for the automatic filtering can be found in the Supporting Information.

## 3 Results

### 3.1 Cohort Creation

The age filter had a precision of 68%, which was calculated by annotating 14, 096 posts, of which 9, 639 actually contained the self-reported age of the user. The inter-annotator agreement was excellent, with a Cohen *κ* of 0.965^45^. After manual verification of the age of the users, and calculating their age in the subreddit, we removed the users not in the expected age range (13 to 25 years old) and ended up with a set of 7, 601 users in the age range of interest.

Of the 7, 601 users, only 760 remained after filtering with the diagnosis regular expression. After manual validation of the 760, only 238 were confirmed as having been diagnosed with SAD (31%). The Cohen *κ* between the annotators was 0.968, which is considered to be excellent agreement^45^.

Similarly, from the 7, 601 users in the age range of interest, only posts from 2, 261 users matched the therapy regular expressions. After manual validation, only 995 users were confirmed as receiving, presently or in the past, therapy for SAD (44%). The Cohen *κ* between the annotators was 0.788, which is considered to be a substantial agreement^45^. For all cases, disagreements between the annotators were manually resolved by the first author. Thus, after manual validation of age (13 to 25) and a confirmed diagnosis or therapy, we identified a set of 1, 102 unique users for our SAD cohort for further analysis. Fig. 2 shows the distribution of the cohort by age and type of SAD confirmation. In the cohort, 131 users report both having a SAD diagnosis **and** having attended therapy. It is worth noting that the Young Adults group aged [19-25] (*n* = 826) is approximately three times the size of the Adolescents group aged [13-18] (*n* = 276).

**Figure 2:**
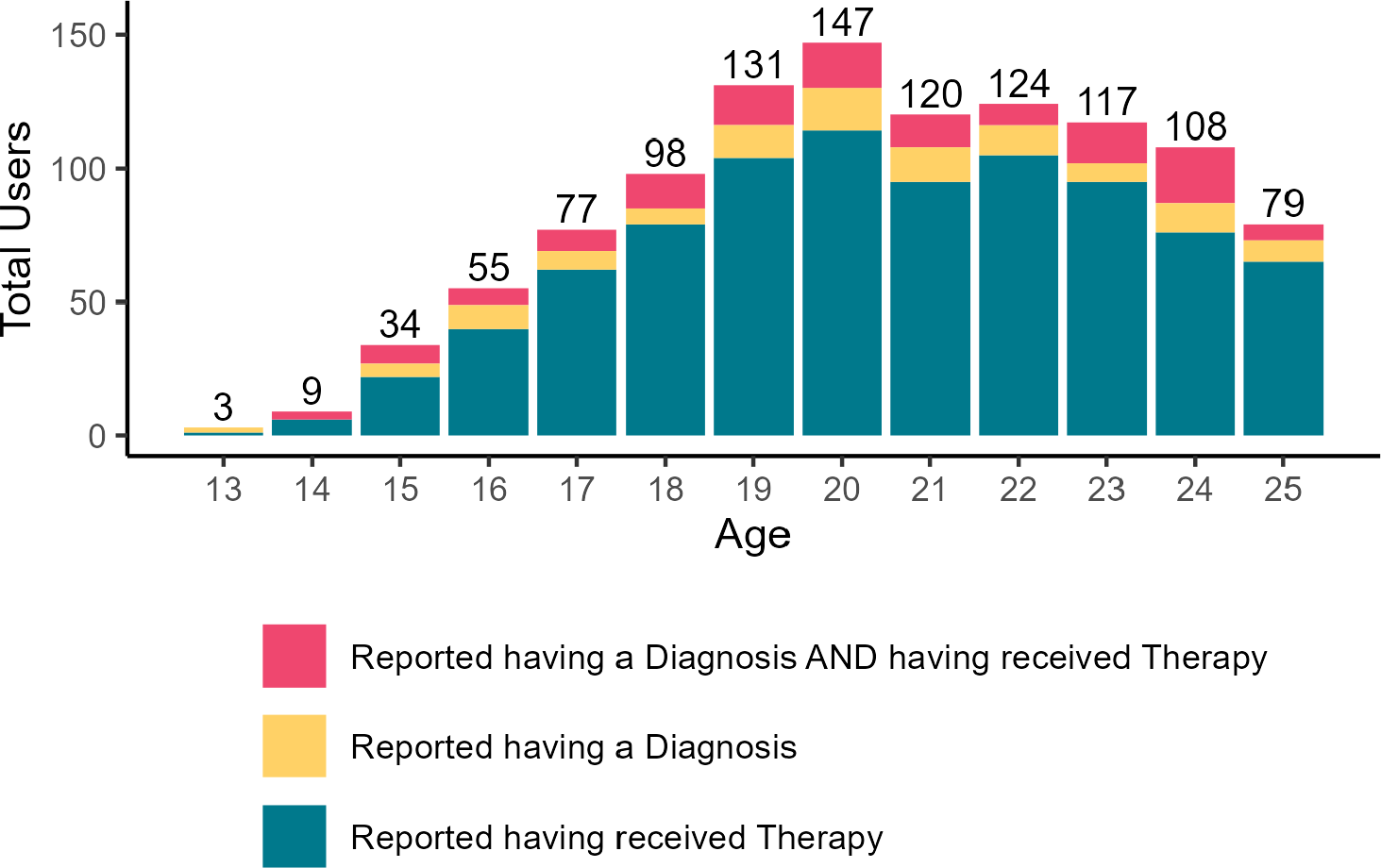
Age distribution within the cohort.

Through a post-annotation review of a random sample of 11% in which the users reported not having a SAD diagnosis (label=‘0’), we found that 61% (*n* = 36) were cases in which the user explicitly mentioned having never been diagnosed, 24% (*n* = 14) discussed other diagnoses, 12% (*n* = 7) were advising other users to get diagnosed, 3% (*n* = 2) were ready or about to be diagnosed with SAD, 3% (*n* = 2) referred to a third person’s SAD diagnosis and 3% (*n* = 2) were diagnosis mentions in a generic context. We also found that 27% (*n* = 16) talked about self-diagnosing with SAD or strongly believed they had it, 7% (*n* = 4) were asking for advice on how to get diagnosed, 5% (*n* = 3) mentioned their parents as barriers to a diagnosis, 5% (*n* = 3) mentioned they never sought a diagnosis, 3%(*n* = 2) mentioned the cost of treatment as a barrier to receiving a diagnosis and one user mentioned COVID-19 as a barrier.

Similarly, in the post-annotation review of a 10% random sample of 124 annotations in which the user did not report having attended therapy for SAD (label=‘0’), we found 15% (*n* = 19) users giving other users advice about therapy, 10% (*n* = 13) asking for advice, 10% (*n* = 13) mentioning they wanted to start therapy, 10% (*n* = 13) sharing information about therapy in a generic manner, 8% (*n* = 10) considering therapy, 8% (*n* = 10) mentioning they were afraid to start or attend therapy, 6% (*n* = 8) mentioning cost as a barrier, 5.6% (*n* = 7) mentioning they were about to start therapy, 5.5% (*n* = 6) considering their parents as a barrier to access treatment, and 4% (*n* = 5) explicitly mentioning they do not want to attend therapy and probably never will.

### 3.2 Case Study: Reported Comorbidities and Substance Abuse Disorders

To show the potential of the generated cohort and associated data, we extracted users’ reports of other comorbidities and mentions of alcohol and/or drug use.

The annotation of reported comorbidities from the cohort shows that *n* = 679 users (61%) report having one or more comorbidities on top of their diagnosis of SAD. The multilabel agreement measured with an unweighted Fleiss’s Kappa^45^ was 0.79 and disagreements between the annotators were manually resolved by the first author.

Table 3 shows the breakdown of the comorbidities reported. The most common was depression (33.21%), followed by general anxiety disorder (6.26%), attention deficit hyperactivity disorder (4.17%), and obsessive-compulsive disorder (2.99%). There were no significant differences in the reporting rate of comorbidities when comparing the age groups or when considering the different levels of certainty in the SAD diagnosis.

**Table 3:**
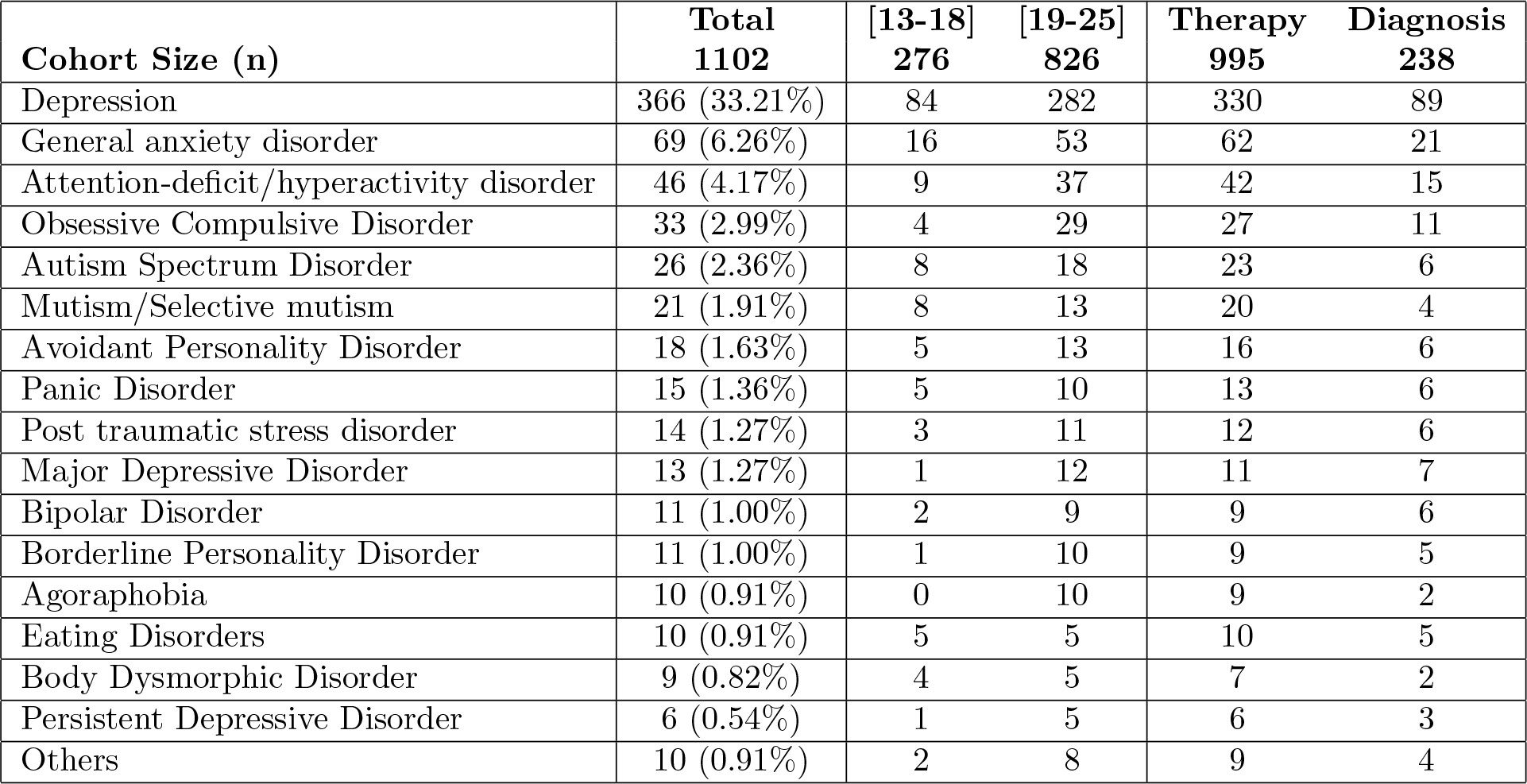
Annotation results of reports of other comorbidities. The table contains the overall totals, the totals in each age group (Adolescents [13-18] and Young Adults [19-25]) and the totals when considering the certainty of a SAD diagnosis (user reports going to therapy or having a SAD diagnosis). Reports of Attention Deficit Disorder are clustered under Attention-deficit/hyperactivity disorder, and Asperger’s under Autism Spectrum Disorder. The “Others” label includes reports of anger problems, cyclothymia, dissociative identity disorder, mood problems, psychosis, psychotic break, schizophrenia, seasonal depression, tic disorder and Tourette Syndrome.

The substance abuse annotation had a multilevel agreement measured with an unweighted Fleiss’s Kappa^45^ of 0.87. Once again, disagreements between the annotators were manually resolved by the first author. Analysis of the disagreements showed that the main problem of the annotation was the difficulty in determining a line in which a reportedly concerning behaviour could be considered addictive behaviour. The labels were revised post-annotation to deal with the subjectivity in the described behaviours related to *concerning* substance use and *addiction* to a substance. Thus, on Table 4 we report 3 types of cases:

**Table 4:** Annotation results of reports of use and abuse of drugs and/or alcohol. The table contains the overall totals for each revised label, the totals in each age group (Adolescents [13-18] and Young Adults [19-25]), and the totals when considering the certainty of a SAD diagnosis (user reports going to therapy or having a SAD diagnosis).

| **Cohort Size (n)** | **Total**<br>**1102** | **[13-18]**<br>**276** | **[19-25]**<br>**826** | **Therapy**<br>**995** | **Diagnosis**<br>**238** |
| --- | --- | --- | --- | --- | --- |
| Concerning Use of Alcohol | 105 (9.53%) | 16 | 89 | 103 | 20 |
| Concerning Use of Drugs | 75 (6.81%) | 15 | 60 | 72 | 13 |
| Concerning Use of Drugs & Alcohol | 31 (2.81%) | 4 | 27 | 30 | 6 |

- Concerning Use of Alcohol: refers to users that describe drinking to cope with SAD, report (now or in the past) having an alcohol addiction problem, or describe a behavior that indicates alcohol addiction. This is the union of the users labeled as having a concerning use of alcohol or alcohol addiction, regardless of their described relationship with drugs.
- Concerning Use of Drugs: refers to users that describe doing recreational drugs to cope with SAD, report (now or in the past) having a drug addiction problem, or describe a behavior that indicates drug addiction. This is the union of the users labeled as having a concerning use of drugs or drug addiction, regardless of their described relationship with alcohol.
- Concerning Use of Drugs & Alcohol: refers to users that match the criteria of the 2 labels above. That is, all users that describe having a concerning behaviour or addiction to alcohol **and** drugs.

We found significant differences (Chi-squared Test p-value=0.02) in the reported rates of alcohol abuse by age group: the Young Adults group had double the frequency of reports (10.77%, *n* = 89) versus the Adolescent group (5.8%, *n* = 16). Interestingly, there was no significant difference in the reported rates of drug abuse when considering the age groups.

## 4 Discussion

Our analysis of a well-defined subset of patients reporting SAD shows that it holds great promise for accessing first-hand reports from a particularly vulnerable population in a costeffective way. There are some limitations to our study which we reflect on in this section.

The low precision of the regular expressions in the creation of the cohort before annotation was due to false positives. Examples of false positives for therapy extraction were mentions of other types of appointments (hair stylists, dentist, etc), sessions (workout, study, etc), treatment (“they treat me badly”, “treatment for ADHD”, etc), exposure therapy, talking about a third party, among others. Further refinement of the regular expressions used would allow for better precision.

As the evaluation of the diagnoses has shown, a large percentage of users in r/SocialAnxiety do not report having a SAD diagnosis or attending therapy at the time they were writing but, considering the long delay in access to treatment, this does not mean the users might not be diagnosed at a later time. The diagnosis discrepancy between the Adolescent group and the Young Adult groups in the cohort confirms the known inverse correlation between access to treatment and age at onset^46,47^ where the earlier a mental disorders manifests, the more delay the patient has in getting treated. While not the focus of this study, this data could provide useful insights on how to improve access to treatment^48^, particularly in underage patients.

The post-annotation analysis of a sample of users that report not having a diagnosis or attending therapy shows that the treatment delay in minors could be due to (1) SAD itself, which limits the patient’s ability to communicate their issues or ask for help, (2) behavior modification (such as avoidance and social isolation) that does not lead to recognition unless or until the symptoms are extreme, (3) normalization of symptoms or the development of coping strategies that interfere with help-seeking during adulthood, (4) the gatekeeping role of parents, who might act as a barrier to mental health services (because it is harder for patients to ask for help directly or the parents’ misconceptions regarding SAD or mental health), or (5) access to child mental health services (cost, geographical availability, lack of information).

Our case study suggests that the reported comorbidities on social media are either consistent or below the incidence rates reported in the literature^49^. For example, the incidence rate reported on social media was 6.26% for General Anxiety Disorder, while it is reported between 0.6 to 27%^49^ in the literature. For ADHD it was 4.17% on social media, while it is reported to be as high as 60% in the published literature^49^. For OCD, we found 2.99%, which is on the low end of the published literature range (2% to 19%^49^). The rate of reported alcohol abuse (9.53%) found on social media is below the published prevalence rates of 16 to 25%^50^. The rate of drug abuse reports found in social media of 6.81% is within the ranges found in the literature (between 2.7 and 39%^50^). Of note, all the comorbidities in the literature for patients diagnosed with SAD vary greatly in reported incidence. This signals a potential inconsistency in traditional studies reflecting shortcomings in this particular patient population that social media listening could partially address.

On the one hand, patients feel freer to express themselves in social media and talk about their condition, comorbidities, and symptoms^51^. Therefore, additional findings could be made from such data in comparison to traditional methods. One the other hand, the detection of diagnoses, treatments, ages, comorbidities and substance abuse problems is limited to what the users report. Thus, the analysis is inherently restricted to what the users chose to disclose in this community specifically dedicated to SAD. Thus, incidence rates on social media could reflect underreporting. It is very likely that many users could have matched the selection criteria but were excluded from the cohort because they did not explicitly report relevant information. That is, they never reported their age or specified having a diagnosis or treatment for SAD. Likewise, it is possible a user had other mental disorder diagnoses and/or substance abuse problems and did not disclose that information. Given that SML is, at its core, passive monitoring, it depends as much as other survey-based observational studies on the patients being truthful. The lower rate of some comorbidities found could also be due to low recall in the methods used.

## 5 Conclusions

In this study we created from social media data a curated cohort of users that report their age and having a diagnosis of SAD, confirming the validity of our approach. We completed a case study to test the potential of the extracted cohort to measure reported comorbidities and substance abuse problems online and found the percentages relatively consistent with the literature. We believe the dataset generated in this study presents multiple opportunities for future research as it could provide insights into the struggles of daily life that the patients face, the barriers they report having to go through to access treatment, and their treatment preferences. This is particularly relevant in underage patients as the posts contain first-hand information about the challenges they face in everyday life, which could aid in removing the barriers patients face when participating in interventional studies. Furthermore, the dataset could allow researchers to tailor the cohort by selecting subsets of patients considering their age, comorbidities, or substance abuse problems, among other possible studies.

## Data Availability

The data that support the findings of this study are available upon reasonable request.
The Reddit data is public and can be found at Academic Torrents.

## 7 Data Availability

The data that support the findings of this study are available upon reasonable request.

The Reddit data is public and can be found at Academic Torrents^44^.

## References

[1] Bogels, S. M. et al. Social anxiety disorder: questions and answers for the DSM-V. Depress. Anxiety 27, 168–189 (2010).

[2] Stein, M. B. & Stein, D. J. Social anxiety disorder. Lancet 371, 1115–1125 (2008).

[3] Stein, D. J. et al. The cross-national epidemiology of social anxiety disorder: Data from the world mental health survey initiative. BMC medicine 15, 1–21 (2017).

[4] Jefferies, P. & Ungar, M. Social anxiety in young people: A prevalence study in seven countries. PloS one 15, e0239133 (2020).

[5] Kessler, R. C. et al. Lifetime prevalence and age-of-onset distributions of dsm-iv disorders in the national comorbidity survey replication. Arch. Gen. Psychiat. 62, 593–602 (2005).

[6] Fehm, L., Beesdo, K., Jacobi, F. & Fiedler, A. Social anxiety disorder above and below the diagnostic threshold: prevalence, comorbidity and impairment in the general population. Soc. Psychiatry Psychiatr. Epidemiol. 43, 257–265 (2008).

[7] Aderka, I. M. et al. Functional impairment in social anxiety disorder. J. Anxiety Disord. 26, 393–400 (2012).

[8] Stein, M. B. An epidemiologic perspective on social anxiety disorder. J. Clin. Psychiat. 67, 3 (2006).

[9] Cruz, E. L. D. d., Martins, P. D. d. C. & Diniz, P. R. B. Factors related to the association of social anxiety disorder and alcohol use among adolescents: a systematic review. J. Pediatr. (Rio J.) 93, 442–451 (2017).

[10] Lydiard, R. B. Social anxiety disorder: Comorbidity and its implications. J. Clin. Psychiatry 62, 17–24 (2001).

[11] Adams, G. C., Balbuena, L., Meng, X. & Asmundson, G. J. When social anxiety and depression go together: A population study of comorbidity and associated consequences. J. Affect. Disord. 206, 48–54 (2016).

[12] Aderka, I. M. et al. Functional impairment in social anxiety disorder. J. of Anxiety Disord. 26, 393–400 (2012).

[13] Levinson, C. A. & Rodebaugh, T. L. Social anxiety and eating disorder comorbidity: The role of negative social evaluation fears. Eat. Behav. 13, 27–35 (2012).

[14] Goodwin, R. D. & Fitzgibbon, M. L. Social anxiety as a barrier to treatment for eating disorders. Int. J. Eat. Disord. 32, 103–106 (2002). URL https://onlinelibrary.wiley.com/doi/10.1002/eat.10051.

[15] Buckner, J. D., Heimberg, R. G., Ecker, A. H. & Vinci, C. A biopsychosocial model of social anxiety and substance use. Depress. Anxiety 30, 276–284 (2013).

[16] Kashdan, T. B. & Herbert, J. D. Social anxiety disorder in childhood and adolescence: Current status and future directions. Clin. Child Fam. Psych. 4, 37–61 (2001).

[17] Erliksson, O. J., Lindner, P. & Mörtberg, E. Measuring associations between social anxiety and use of different types of social media using the swedish social anxiety scale for social media users: A psychometric evaluation and cross-sectional study. Scand. J. Psychol. 61, 819–826 (2020).

[18] Ricky, C., Nawaf, M. et al. Factors associated with delayed diagnosis of mood and/or anxiety disorders. Health. Promot. Chronic Dis. Prev. Can. 37, 137 (2017).

[19] Olteanu, A., Castillo, C., Diaz, F. & Kıcıman, E. Social data: Biases, methodological pitfalls, and ethical boundaries. Front. Big Data 2, 13 (2019).

[20] Glazier, B. L. & Alden, L. E. Social anxiety and biased recall of positive information: It’s not the content, it’s the valence. Behav. Ther. 48, 533–543 (2017).

[21] Rozin, P. & Royzman, E. B. Negativity bias, negativity dominance, and contagion. Pers. Soc. Psychol. Rev. 5, 296–320 (2001).

[22] Brozovich, F. & Heimberg, R. G. An analysis of post-event processing in social anxiety disorder. Clin. Psychol. Rev. 28, 891–903 (2008).

[23] Wright, D. B., London, K. & Waechter, M. Social anxiety moderates memory conformity in adolescents. Appl. Cognitive Psychol. 24, 1034–1045 (2010).

[24] Jong, S. T., Stevenson, R., Winpenny, E. M., Corder, K. & Van Sluijs, E. M. F. Recruitment and retention into longitudinal health research from an adolescent perspective: a qualitative study. BMC. Med. Res. Methodol. 23, 16 (2023).

[25] Parrish, D. E., Duron, J. F. & Oxhandler, H. K. Adolescent recruitment strategies: lessons learned from a university-based study of social anxiety. Soc. Work Res. 41, 213–220 (2017).

[26] Teague, S. et al. Retention strategies in longitudinal cohort studies: a systematic review and meta-analysis. BMC Med. Res. Methodol. 18, 1–22 (2018).

[27] O’Day, E. B. & Heimberg, R. G. Social media use, social anxiety, and loneliness: A systematic review. Comput. Hum. Behav. Rep. 3, 100070 (2021).

[28] Reid, D. J. & Reid, F. J. Text or talk? social anxiety, loneliness, and divergent preferences for cell phone use. Cyberpsychol. Behav. 10, 424–435 (2007).

[29] Schmidt, A. L., Rodriguez-Esteban, R., Gottowik, J. & Leddin, M. Applications of quantitative social media listening to patient-centric drug development. Drug Discov. Today 27, 1523–1530 (2022).

[30] Wongkoblap, A., Vadillo, M. A. & Curcin, V. Researching mental health disorders in the era of social media: Systematic review. J. Med. Internet Res. 19, e228 (2017).

[31] Chancellor, S. & De Choudhury, M. Methods in predictive techniques for mental health status on social media: a critical review. NPJ Digit. Med. 3, 43 (2020).

[32] Su, C., Xu, Z., Pathak, J. & Wang, F. Deep learning in mental health outcome research: a scoping review. Transl. Psychiatry 10, 116 (2020).

[33] Coppersmith, G., Dredze, M., Harman, C. & Hollingshead, K. From adhd to sad: Analyzing the language of mental health on twitter through self-reported diagnoses. In Proceedings of the 2nd Workshop on Computational Linguistics and Clinical Psychology: From Linguistic Signal to Clinical Reality, 1–10 (Association for Computational Linguistics, 2015).

[34] Saha, K., Yousuf, A., Boyd, R. L., Pennebaker, J. W. & De Choudhury, M. Social media discussions predict mental health consultations on college campuses. Sci. Rep. 12, 123 (2022).

[35] Reece, A. G. & Danforth, C. M. Instagram photos reveal predictive markers of depression. EPJ Data Sci. 6, 15 (2017).

[36] Guntuku, S. C., Yaden, D. B., Kern, M. L., Ungar, L. H. & Eichstaedt, J. C. Detecting depression and mental illness on social media: an integrative review. Curr. Opin. Behav. Sci. 18, 43–49 (2017).

[37] Won, H.-H. et al. Predicting national suicide numbers with social media data. PLoS ONE 8, e61809 (2013).

[38] Robinson, J. et al. Social media and suicide prevention: a systematic review. Early Interv. Psychiatry 10, 103–121 (2016).

[39] Guntuku, S. C., Ramsay, J. R., Merchant, R. M. & Ungar, L. H. Language of adhd in adults on social media. J. Atten. Disord. 23, 1475–1485 (2019).

[40] Shen, J. H. & Rudzicz, F. Detecting anxiety through reddit. In Proceedings of the Fourth Workshop on Computational Linguistics and Clinical Psychology — From Linguistic Signal to Clinical Reality, 58–65 (Association for Computational Linguistics, 2017). URL http://aclweb.org/anthology/W17-3107.

[41] Stemmer, M., Parmet, Y. & Ravid, G. Identifying patients with inflammatory bowel disease on twitter and learning from their personal experience: Retrospective cohort study. Journal of medical Internet research 24, e29186 (2022).

[42] Sarker, A. et al. Discovering cohorts of pregnant women from social media for safety surveillance and analysis. Journal of medical Internet research 19, e361 (2017).

[43] Baumgartner, J., Zannettou, S., Keegan, B., Squire, M. & Blackburn, J. The pushshift reddit dataset. In Proceedings of the international AAAI conference on web and social media, vol. 14, 830–839 (2020).

[44] [stuck in the matrix & Watchful1]. Reddit comments/submissions 2005-06 to 2022-12. https://academictorrents.com/details/7c0645c94321311bb05bd879ddee4d0eba08aaee (2023). [Online; accessed 12-September-2023].

[45] Fleiss, J. L., Levin, B., Paik, M. C. et al. The measurement of interrater agreement. Statistical methods for rates and proportions 2, 22–23 (1981).

[46] Christiana, J. et al. Duration between onset and time of obtaining initial treatment among people with anxiety and mood disorders: an international survey of members of mental health patient advocate groups. Psychol. Med. 30, 693–703 (2000).

[47] Wang, P. S. et al. Failure and delay in initial treatment contact after first onset of mental disorders in the national comorbidity survey replication. Arch. Gen. Psychiatry 62, 603–613 (2005).

[48] Olfson, M. Barriers to the treatment of social anxiety. Am. J. Psychiatry 157, 521–527 (2000).

[49] Koyuncu, A., İnce, E., Ertekin, E. & Tükel, R. Comorbidity in social anxiety disorder: diagnostic and therapeutic challenges. DIC 8, 1–13 (2019).

[50] Bakken, K., Landheim, A. & Vaglum, P. Substance-dependent patients with and without social anxiety disorder: occurrence and clinical differences: a study of a consecutive sample of alcohol-dependent and poly-substance-dependent patients treated in two counties in norway. Drug and alcohol dependence 80, 321–328 (2005).

[51] Humphrey, L. et al. A comparison of three methods to generate a conceptual understanding of a disease based on the patients’ perspective. Journal of patient-reported outcomes 1, 1–12 (2017).

