## Supporting Information for "Studying social anxiety without triggering it: Establishing an age-controlled cohort of social media users for observational studies"

December 11, 2023

#### 1 Regular Expressions

Here the regular expressions used at the different stages can be found.

##### 1.1 Regular Expressions for Age detection

Posts that matched the regular expression *re.age* were kept and then matched against the regular expressions listed in *re.time*, *re.dosage*, *re.money*, *re.percentage*, *re.other.nrs*, *re.common* and *re.fraction*. Posts that matched the condition were then manually annotated as described in the paper.

```
re.age = "\\b(1[2-9]|2[0-5])(\\b|f|m)"
re.time = c("second(s)?", "minute(s)?", "hour(s)?", "day(s)?", "
    week(s)?", "month(s)?", "hr(s)?", "min(s|.)?", "sec(s|.)?")
re.dosage = c("drop(s)?", "pill(s)?", "time(s)?", "mg(.)?", "ml(.)
    ?", "oz(.)?", "vial(s)?", "capsule(s)?", "dose", "mcg(.)?", "mm
    (.)?")
re.money = c("£", "€", "\\$", "pound(s)?", "dollar(s)?", "euro(s)
    ?")
re.percentage = c("%", "percent")
```

```
re.other.nrs = c("24(/|-)7", "20(/|-)20", "catch( |-)?22", "
    patient(s)?", "covid(-| )?19", "people", "page(s)?", "degree(s)
    ?", "message(s)?", "session(s)?")
re.common = c("was \\d\\d|\\d\\d years ago", "you('re| are| r) (
    only)? \\d\\d")
re.fraction = c"(\\d)?\\d(\\.|,|:)\\d(\\d)?", "\\d\\d/\\d\\d", "\\
    d\\d-\\d\\d")
```

#### 1.2 Regular Expressions for Diagnosis Confirmation

Sentences that matched the regular expression *re.diagnosis* or *re.therapy* were kept, group by author and sorted chronologically for manual annotation.

```
re.diagnosis = "diagno"
re.therapy = c("therapy", "counseling", "session", "therapist", "
    doctor", "\\bappointment", "treat(ed|ment)", "psychologist", "
    psychotherapist", "shrink",  "psychoanalyst", "clinician")
```

#### 1.3 Regular Expressions for Detection of Comorbidities

Sentences that matched the regular expressions listed in *re.depression*, *re.gad*, *re.ocd*, *re.adhd*, *re.avpd*, *re.bpd*, *re.panic*, *re.autism*, *re.bipolar*, *re.ptsd*, *re.mutism*, *re.ed* and *re.other.disorder*, were kept, group by author and sorted chronologically for manual annotation.

```
re.depression = c("(major |persistent )?depressive disorder", "
    depression", "\\bMDD\\b", "dysthymia")
re.gad = c("\\bGAD\\b", "general(i(s|z)ed)? anxiety( disorder)?")
re.ocd = c("\\bOCD\\b", "obsessive compulsive disorder")
```

```
re.adhd = c("\\bADHD\\b", "attention deficit hyperactivity
    disorder")
re.avpd = c("\\bAVPD\\b", "avoidant personality( disoder)?")
re.bpd = c("\\bBPD\\b", "borderline( personality( disorder)?)?")
re.panic = c("panic disorder")
re.autism = c("autism( spectrum( disorder)?)?", "\\bASD\\b", "
    autistic",)
re.bipolar = c("bipolar( disorder)?")
re.ptsd = c("\\bPTSD\\b", "post traumatic stress disorder")
re.mutism = c("(selective )?mutism")
re.ed = c("\\bED\\b", "eating disorder", "anorexi(a|c)", "bulimi(a
    |c)")
re.other.disorder  = c("epilepsy", "asperger(s)?", "psychotic
    break", "body dysmorphi(a|c)( disorder)?", "\\bADD\\b", "
    attention deficit( disorder)?",  "tic disorder", "agoraphobi(a|
    c)", "schizophreni(a|c)", "dissociative identity disorder")
```

##### 1.4 Regular Expressions for Substance Abuse

Sentences that matched the regular expressions listed in *re.alcohol*, *re.drugs*, *re.smoking* and *re.generic.substance.abuse* were kept, group by author and sorted chronologically for manual annotation.

```
re.alcohol = c("alcohol(ic|ism)?", "drinking problem", "drunk", "
    drinking", "buzzed", "tipsy")
```

```
re.drugs = c("drug(s)?", "\\bTHC\\b", "\\bCBD\\b", "\\bhemp\\b", "
    cannabis", "marijuana", "ayahuasca", "\\bweed", "cocaine", "\\
    bcoke\\b", "\\bcrack\\b", "hallucinogen(s)?", "heroin", "\\bLSD
    \\b", "\\bacid\\b", "mescaline", "peyote", "\\bPCP\\b", "angel
    dust", "psilocybin", "salvia", "magic mushroom(s)?", "\\
    bshrooms\\b", "khat", "kratom", "ecstasy", "\\bmolly\\b", "\\
    bmdma\\b", "methamphetamine", "\\bmeth\\b", "\\bMDMA\\b", "\\
    bDMT\\b")

re.smoking = c("smok(e|ing|es)", "tobacco", "nicotine", "\\bvap(e|
    ing|es)", "cigarette" )

re.generic.substance.abuse = c("addict(ion|ed|ive)?", "rehab", "
    overdose", "withdrawal")
```

### 2 Annotation Guidelines

The following subsections have the annoation guidelines as they were given to the annotators.

#### 2.1 Annotation Guidelines for Age

The data was extracted from the subreddit r/socialanxiety. The dataset consists of 14,027 posts that match a series of regular expressions to detect 2 digits numbers in the [12-25] range and don't match a set of regular expressions that indicate the number is unlikely to be the age of the user. Each row corresponds to a post and contains a unique identifier, all the text (title + content if it's the first post of a thread or just content if it's a comment), the sentences that have a 2 digit number in the correct range and a empty column 'age' where the annotation should be added. The labels to annotate are:

- The numeric age for the cases where the user reports their age (first person report).
- NA when no age is reported, the reported age is not that of the user who wrote the post or the age is unclear or lacks specificity.

The annotation should be done based on the numbered sentences, unless the annotator believes more information is required, in which case they can also use the complete post column to get more context.

**Positive age report:**

1. The user explicitly states their age. Examples:
   - "I'm 20 years old" →age = 20
   - "I turned 19 last week" →age = 19
2. The user reports their age following the standard Reddit format (age/gender) to mention their demographics, that is, their age following a letter to indicate their gender. Examples:
   - "I'm 25F..." →age = 25
   - "Me 24M..." →age = 24
3. The age is easily calculated from the sentence. Examples:
   - "My 15th birthday is in 2 months" →age = 14
   - "It started when I was 17, two years ago" →age = 19
   - "I'm almost 20" →age = 19

**Negative age report:**

1. The user does not report their age in the post.

2. The user reports their age in an ambiguous manner (“I'm a teenager”, “I'm in my early twenties”, “I just started high school”).

3. There is a reported age in the post but it's not the age of the user, that is, it refers to someone else (“My son 14M”, “She's just 20 years old”, “You are only 15 yo”)

4. All other cases where the age is unclear or not present.

#### 2.2 Annotation Guidelines for Diagnosis

The data was extracted from the subreddit r/socialanxiety. The dataset consists of a user aggregation of all the sentences that match a regular expression to detect diagnosis mentions. Each row corresponds to a user and contains a unique identifier, all the sentences that match the regular expression (sorted chronologically) separated by a semicolon and an empty column ‘diagnosis’ where the annotation should be added.

For each user the annotator must look at the sentences and decide on the label:

- ‘1’: the user reports having a Social Anxiety Disorder (SAD) diagnosis or having been diagnosed with SAD.

- ‘0’ the user reports not having or wanting a diagnosis, is asking about other people's diagnosis or information related to it, or mentions being self-diagnosed, is talking about a diagnosis that is not SAD, or any other case where the user does not indicate having a diagnosis.

As each row contains all the sentences written by the user that match the regular expression in chronological order, it's possible that the status of the user diagnosis contradicts itself throughout the sentences, e.g. the first reports having no diagnosis or being self-diagnosed and later reports having a diagnosis. Once a user mentions having a diagnosis the final label will be positive (‘1’) for all cases, except those when the user reports afterward they were misdiagnosed.

On Reddit social anxiety disorder is also called a variety of names, the most common being "social anxiety", "SA", "SAD", "social phobia" and in some cases just "anxiety". Other disorders might also be mentioned and even though they are not relevant for this annotation it's worth taking a look at their abbreviations to avoid confusion and save time:

- MDD - major depressive disorder
- PDD - persistent depressive disorder
- GAD - general anxiety disorder
- ADHD - attention deficit hyperactivity disorder
- OCD - obsessive compulsive disorder
- ASD - autism spectrum disorder
- AVPD - avoidant personality disorder
- ADD - attention deficit disorder
- PTSD - post traumatic stress disorder
- BPD - borderline personality disorder
- ED - eating disorder
- DID - dissociative identity disorder

**Positive diagnosis report (diagnosis='1')**

1. The user explicitly reports having a diagnosis of SAD. Examples: "I've been diagnosed with SA", "My therapists diagnosed me with social phobia", "When I was 15 I was diagnosed with SAD".

2. The user reports having a diagnosis but the disease is not mentioned. Examples: “Yes, I was diagnosed 2 months ago”, “I have a diagnosis”, “My therapists confirmed I have it”.

3. The user reports their doctor (therapist/psychologist/etc) agrees or believes they have SAD. Example: “My therapist agreed with my self-diagnosis”.

4. Any other case where the annotator believes the user is talking about their SAD diagnosis.

**Negative diagnosis report (diagnosis=‘0’)**

1. The user reports not having a diagnosis of SAD. Examples: “I’ve never been diagnosed”, “I can’t afford to get diagnosed”, “I don’t want a diagnosis”, “I don’t know how to go about getting diagnosed”.

2. The user reports having a diagnosis of another disease/disorder but no mention of SAD. Examples: “I’ve been diagnosed with depression”, “I was diagnosed with BPD last year”,

3. The user reports being self-diagnosed. Examples: “I’m self-diagnosed”, “I did self diagnose with SA ”,

4. The user reports having social anxiety but makes no mention of a diagnosis or treatment. Example: “I was just recently diagnosed with ADHD-PI. [...] The combination of SA and ADHD sucks. Even when I really want to, I find it hard to focus on what people are saying. I get bits and pieces of what people are saying and it’s frustrating and makes my SA worse”,

5. All other cases where it’s clear the user doesn’t have a diagnosis.

#### 2.3 Annotation Guidelines for Therapy

The data was extracted from the subreddit r/socialanxiety. The dataset consists of a user aggregation of all the sentences that match a regular expression to detect therapy related sentences (mentions of therapy, sessions, doctor, psychologist, therapists, etc). Each row corresponds to a user and contains a unique identifier, all the sentences separated by a semicolon and an empty column 'therapy' where the annotation should be added.

The labels to annotate are:

- '1': the user reports going or having gone to therapy in the past for Social Anxiety Disorder (SAD). SAD does not necessarily have to be explicitly mentioned.

- '0' the user reports wanting to go to or try therapy, is asking for information about therapy, mentions going to therapy for a specific reason not related to SAD, or any other reason where they mention therapy but it's not in the context of them getting treatment.

As each row contains all the sentences written by the user that match the regular expression in chronological order, it's possible that the status of the user therapy contradicts itself throughout the sentences, e.g. the first reports they never tried therapy or they are about to start therapy and later reports going to therapy. Once a user mentions having gone to therapy, the final label will be positive ('1') for all cases.

On Reddit social anxiety disorder is also called a variety of names, the most common being "social anxiety", "SA", "SAD", "social phobia" and in some cases just "anxiety". Other disorders might also be mentioned and even though they are not relevant for this annotation it's worth taking a look at their abbreviations to avoid confusion and save time:

- MDD - major depressive disorder

- PDD - persistent depressive disorder

- GAD - general anxiety disorder

- ADHD - attention deficit hyperactivity disorder
- OCD - obsessive compulsive disorder
- ASD - autism spectrum disorder
- AVPD - avoidant personality disorder
- ADD - attention deficit disorder
- PTSD - post traumatic stress disorder
- BPD - borderline personality disorder
- ED - eating disorder
- DID - dissociative identity disorder

**Positive therapy report (therapy = '1')**

1. The user reports that they are currently attending therapy or currently have a therapist. SAD does not necessarily have to be explicitly stated. Examples: "I've only gone to a few sessions so far and my SA. . . ", "I talked with my therapist about [...]", "I just started therapy", "I'm thinking about finding a new therapist".

2. The user reports that they used to have a therapist or used to go to therapy. Again, the disorder does not necessarily have to be explicitly stated. Examples: "I was on therapy to help my sad for a long time", "I think I'm going to start therapy again", "Yes, I did 12 sessions because that's all my insurance covered".

3. The user reports going to group therapy. Examples: "I started group therapy a few weeks ago", "I think group therapy really helped my SA".

4. The user reports going to a doctor or talking to a doctor about SAD. Examples: "I eventually found a doctor I felt a good connection with so I felt comfortable sharing."

**Negative therapy report (therapy = '0')**

1. The user reports doing therapy with no professional intervention. Examples: "I've been doing exposure therapy by myself", "I've been doing self counseling".

2. The user reports going to a doctor but not necessarily for SAD. Example: "I only go out for necessary interactions like going to the doctor", "My mom used to make me go to counseling after my parents got a divorce"

3. The user reports they are about to start therapy. Example: "My first therapy session is tomorrow", "I have my first appointment with my psychologist in a month"

4. The user reports going to therapy for a specific reason unrelated to SAD. Example: "I went to therapy when my parents divorced", "I was diagnosed with BPD a year ago and have been going to therapy weekly since."

5. All other cases where it's clear the user is not talking about themselves attending therapy: they are discussing cost, asking other users for information or advice, talking about the barriers they face to get therapy, etc.

#### 2.4 Annotation Guidelines for Substance Abuse

The data was extracted from the subreddit r/SocialAnxiety. The dataset consists of a user aggregation of all the sentences that match a regular expression to detect substance abuse related sentences (mentions of drinking, smoking, doing drugs, overdose, etc). Each row corresponds to a user and contains a unique identifier, all the sentences separated by a semicolon and an empty column 'substance abuse' where the annotation should be added.

To facilitate the annotation a dropdown menu has been added in the 'substance abuse' column which allows the easy selection of the appropriate label. However, there are cases where more than one label could be annotated. As some combinations of labels are more likely, we introduced 2 to simplify those few combined labels: 'Concerning use of A&D' and

'Addiction - A&D'. In other multiple label cases, the annotator should select one appropriate label from the dropdown menu and annotate the rest in the 'Notes' column. The labels to annotate are:

- 'Not concerning': the user reports having tried alcohol and or drugs in the past, describing a normal drinking behavior or a normal recreational use of legal drugs. Examples: "I'm quiet, reserved, and hate parties and alcohol"; "I smoke a lot and have pretty bad SA, smoking can be really fun"; "I've tried everything from decaffinated tea, propranolol, CBD, deep breaths, yoga, etc etc."; "Weed is the only one that works, but only while I'm high, and I'm not going to be high 24/7"

- 'Concerning use of alcohol': the user reports drinking to cope with SAD, but does not describe a behavior that seems to indicate addiction. Examples: "I love going out [...] but I can only enjoy it when I'm drunk"; "Doing my presentation drunk because this fucking university can't just do essays and leave it be"

- 'Concerning use of drugs': the user reports doing recreational drugs to cope with SAD, but does not describe a behavior that seems to indicate addiction. Examples: "I need weed to interact with people"; "weed helps me give presentations at school"

- 'Addiction - Alcohol': the user reports having an addiction problem with alcohol or having had an addiction problem in the past, or describes a behavior that indicates addiction. Examples: "I find myself drinking alcohol before going to work to numb down my anxiety"; "I'm an alcoholic"; "I use to have an alcohol problem".

- 'Addiction - Drugs': the user reports having an addiction problem to drugs or having had an addiction problem in the past, or describes a behavior that indicates addiction. Examples: "I'm 20 with a history of drug abuse", "i got addicted to an opiate called kratom but bounced back and recovered"; "I struggled and still do struggle with substance abuse, mainly opiates and various other pills"

- ‘NA’: the user talks about alcohol and/or drugs but it has nothing to do with their relationship to drugs and alcohol (they talk about other people, hypothetical, etc). Examples: “I know that cannabis can have a lot of calming and healing properties and I am really curious to see if it will work for me”, “have you tried alcohol?”; “Drunk dad called my workplace, caused a lot of trouble for me?.”
- ’Other’: the user reports having other kinds of addictions. The details should be added on the Notes.

#### 2.5 Annotation Guidelines for Other Diagnosis

The data was extracted from the subreddit r/SocialAnxiety. The dataset consists of a user aggregation of all the sentences that match a regular expression to detect other mental disorders. Each row corresponds to a user and contains a unique identifier, all the sentences separated by a semicolon, an empty column ‘comorbidities’ where the annotations should be added and a ‘Notes’ column. To facilitate the annotation a dropdown menu has been added in the ‘comorbidities’ column which allows the easy selection of the appropriate label. However, there are cases where more than one comorbidity will be reported by a user. In those cases, the annotator should select one appropriate label from the dropdown menu and annotate the rest of the comorbidities present in the text in the ‘Notes’ column. Other mental disorders reported by the user that are not present in our label set should also be annotated as ‘Other’ and then written down in the ‘Notes’ column. Finally, it’s worth clarifying that hypothetical or theoretical self-diagnoses are not considered a reported comorbidity (i.e. “I think I have PTSD”, “I suspect I’m autistic”, “I self-diagnosed with avpd”). The labels to annotate are:

- ’MDD - major depressive disorder’: the user reports having major depressive disorder.
- ’PDD - persistent depressive disorder’ : the user reports having persistent depressive disorder.

- ’D - depression’: the user reports having depression or being depressed.
- ’GAD - general anxiety disorder’: the user reports having general anxiety disorder.
- ’ADHD - attention deficit hyperactivity disorder’: the user reports having attention deficit hyperactivity disorder.
- ’OCD - obsessive compulsive disorder’: the user reports having obsessive compulsive disorder.
- ’ASD - autism/Asperger’s disorder’: the user reports being on the autism spectrum or having Asperger’s.
- ’AVPD - avoidant personality disorder’: the user reports having avoidant personality disorder.
- ’ADD - attention deficit disorder’: the user reports having attention deficit disorder.
- ’PTSD - post traumatic stress disorder’: the user reports having post traumatic stress disorder.
- ’BPD - borderline personality disorder’: the user reports having borderline personality disorder.
- ’ED - eating disorder’: the user reports having an eating disorder.
- ’DID - dissociative identity disorder’: the user reports having dissociative identity disorder.
- ’BP - bipolar disorder’: the user reports being bipolar.
- ’AGO - agoraphobia’: the user reports having agoraphobia.
- ’NA’: the user does not report having any other comorbidities besides SAD. Examples: “it makes me look like I’m mentally disabled or autistic which makes me more anxious”, “Do you happen to have autism?”.

- 'Other': Any other mental disorder reported by the user and not in the current list should then be annotated on the Notes column.
